# Supporting COVID-19 policy response with large-scale mobility-based modeling

**DOI:** 10.1101/2021.03.20.21254022

**Authors:** Serina Chang, Mandy L. Wilson, Bryan Lewis, Zakaria Mehrab, Komal K. Dudakiya, Emma Pierson, Pang Wei Koh, Jaline Gerardin, Beth Redbird, David Grusky, Madhav Marathe, Jure Leskovec

## Abstract

Social distancing measures, such as restricting occupancy at venues, have been a primary intervention for controlling the spread of COVID-19. However, these mobility restrictions place a significant economic burden on individuals and businesses. To balance these competing demands, policymakers need analytical tools to assess the costs and benefits of different mobility reduction measures.In this paper, we present our work motivated by our interactions with the Virginia Department of Health on a decision-support tool that utilizes large-scale data and epidemiological modeling to quantify the impact of changes in mobility on infection rates. Our model captures the spread of COVID-19 by using a fine-grained, dynamic mobility network that encodes the hourly movements of people from neighborhoods to individual places, with over 3 billion hourly edges. By perturbing the mobility network, we can simulate a wide variety of reopening plans and forecast their impact in terms of new infections and the loss in visits per sector. To deploy this model in practice, we built a robust computational infrastructure to support running millions of model realizations, and we worked with policymakers to develop an intuitive dashboard interface that communicates our model’s predictions for thousands of potential policies, tailored to their jurisdiction. The resulting decision-support environment provides policymakers with much-needed analytical machinery to assess the tradeoffs between future infections and mobility restrictions.

## 1 INTRODUCTION

The COVID-19 pandemic has wreaked havoc on lives and livelihoods across the globe. In an effort to contain the virus, policy-makers have turned to non-pharmaceutical interventions, such as restricting mobility, in order to limit contact and reduce disease transmission between individuals. To this end, many US states and local governments have closed or required reduced occupancy at places such as restaurants and gyms [7]. However, these measures come at a heavy cost to individuals and businesses: for example, over 160,000 US businesses closed due to COVID-19 between March and August 2020 [36].

The next few months will continue to pose challenges to public health and economic activity. It is imperative during this time to provide policymakers with analytical tools that can help them develop optimal solutions to minimize new COVID-19 infections while also minimizing damage to businesses. They need a tool that can quantitatively assess, in near real-time, the tradeoffs between mobility and new infections. Furthermore, this tool should be fine-grained, able to test out heterogeneous plans—for example, allowing one level of mobility at essential retail, another level at retail, and yet another at restaurants—so that policymakers can tailor restrictions to the specific risks and needs of each sector. Despite this granularity, the tool also needs to be scalable, supporting analyses for an exponential number of potential policies so that policymakers can select the best option among them for their jurisdiction.

To fullfill these needs, we present a decision-support tool, which we built based on interactions with the Virginia Department of Health (VDH) to support their decision-making on mobility reduction policies. Our approach begins with our state-of-the-art epidemiological model [8], which integrates large-scale mobility and mask-wearing data to accurately capture the spread of COVID-19. Our model overlays transmission dynamics on a time-varying mobility network which encodes the hourly movements of individuals from neighborhoods to specific points of interest (POIs), such as restaurants or grocery stores. Since we model infections in tandem with mobility, our model can provide the multifaceted analyses necessary to understand the costs and benefits of a policy; for example, by quantifying predicted infections and the number of POI visits lost per sector, which can serve as a proxy for economic impact. By design, our model is ne-grained, as it simulates who is getting infected where and when down to the individual POI and hour. Our model is also flexible, since we can modify any one of its inputs—for example, modifying mobility for a subset of POIs to reflect a change in policy, or modifying transmission rates per neighborhood to indicate vaccination effects—and straightforwardly run the model with the new inputs to observe the effects of the hypothetical change.

Finally, to scale our model, we build a robust infrastructure to handle computational challenges. The mobility networks that we model are large, with billions of hourly edges between POIs and neighborhoods. Furthermore, the flexibility of our approach—for example, being able to simulate different POI categories at different levels of mobility—results in an exponential number of hypothetical scenarios to test. In order to support predictions for thousands of scenarios, we design a lightweight, vectorized implementation of our model, and build a large-scale system to run hundreds of models in parallel.^1^ By leveraging this system, we are able to compress 2 years of compute time into the span of a few days.

### Advances in the current work

In building this tool, we have greatly extended our epidemiological model since its original implementation [8]. We have introduced new features including (1) variation in mask-wearing over time; (2) a time-varying base transmission rate; (3) a time-varying death detection rate; and (4) model initialization based on historical reported deaths. These additions allow us to accurately fit daily deaths in Virginia, and we also show that the inclusion of our new features contribute substantially toward reducing model loss (Section 3.1). Furthermore, whereas our original work focused on the first two months of the pandemic (March and April 2020), in this work we focus on recent months (November 2020 to January 2021), which are more relevant to current policy-making. We have also fitted the model on new, smaller metropolitan areas in Virginia. Importantly, the experimental results in this work are consistent with and extend our original analyses, showing that the high-level findings from our first work generalize to new time periods and smaller regions. For example, we continue to find that mobility patterns are predictive of disparities in infection rates between lower- and higher-income neighborhoods, and that certain POI categories like restaurants are far more dangerous to fully reopen than others (Section 3.2). Finally, to create a finished product that policymakers can directly use, we developed a new dashboard that can communicate thousands of results from our model. Our resulting interface includes multiple interactive panels, where policymakers can select various proposed changes in mobility, and observe how these changes would affect predicted infections over time and losses in POI visits (Section 3.3).

### Supporting public health decision-making

Our group has been supporting various federal, state and local public health authorities for over a year now as they respond to the pandemic. The need for such a tool became clear to us during the course of sustained response efforts. This tool was designed to fulfill public health officials’ desire to have a quantitative and comprehensive analysis of a range of reopening policies. VDH reviewed a prototype of the tool and provided valuable feedback on how best to present the data to maximize clarity and applicability from a public health perspective. This guidance was integral to the final design of the dashboard presented in this paper. While we focus on the state of Virginia for illustrative purposes, the tool can be generalized and used in other states as well.

## 2 OUR APPROACH

In this section, we break down our approach: the datasets that we use (Section 2.1), our epidemiological model (Section 2.2), and the computational infrastructure that we developed to produce predictions at scale (Section 2.3). Figure 1 also provides a summary of our process, illustrating the main steps of our approach and where different data sources are integrated along the way.

**Figure 1.**
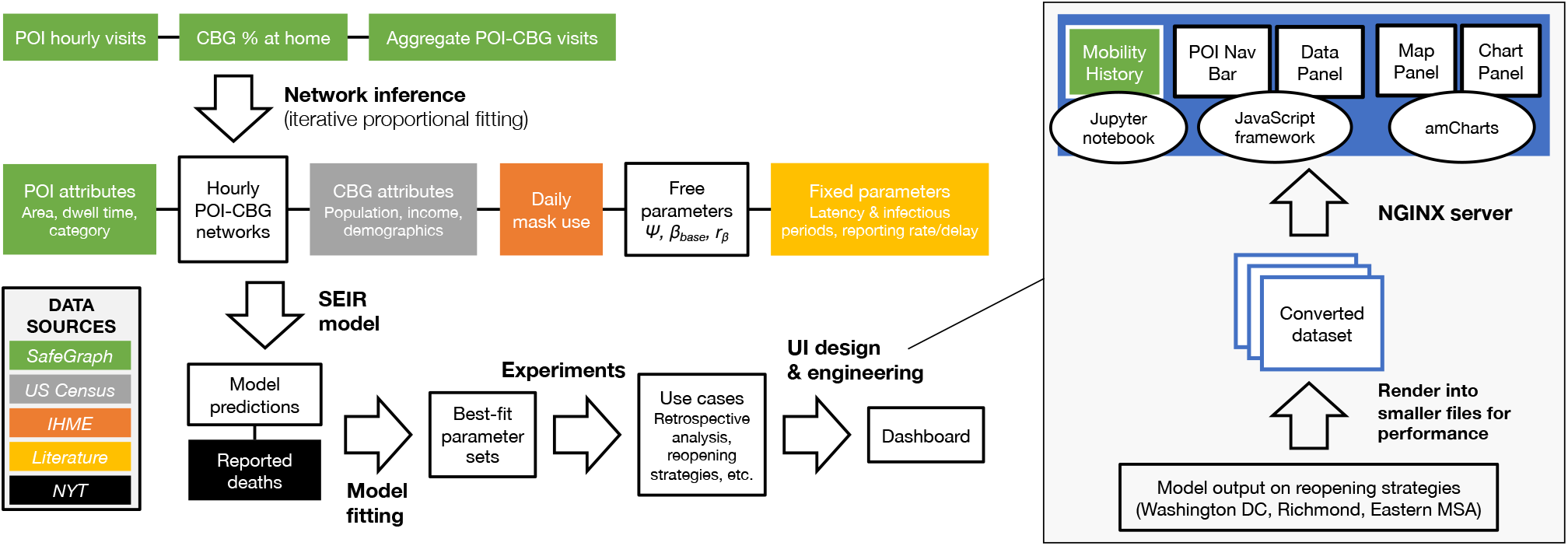
Illustration of our approach. We integrate data from many sources to run, evaluate, and analyze our model. We build a computational infrastructure to support millions of model realizations during the model fitting and experiment steps of our pipeline. Our UI design enables a smooth experience, so that policymakers can easily compare different reopening policies.

### 2.1 Large-scale data

#### Fine-grained mobility data (SafeGraph)

Mobility data capture important changes in population behavior over time: for example, in Figure 2, we see that mobility fell dramatically in March 2020, then slowly climbed back up during the following months until receding again near the end of the year. These patterns reflect where and when individuals may have been coming into contact with one another, and thus inform our understanding of transmission risks and how to mitigate them. We use data from SafeGraph, a company that anonymizes and aggregates location data from mobile apps. SafeGraph’s Places^2^ and Weekly Patterns^3^ datasets provide detailed information about millions of points of interest (POIs), which are non-residential locations that people can visit. For each POI, SafeGraph provides estimates of its hourly visit counts, as well as weekly estimates of which census block groups the visitors are coming from. In addition, SafeGraph provides each POI’s North American industry classification systems (NAICS) category, its physical area in square feet, and its median visit duration in minutes (the “dwell time”). We also use SafeGraph’s Social Distancing Metrics^4^ dataset, which contains daily estimates of the proportion of people staying at home in each CBG.

**Figure 2.**
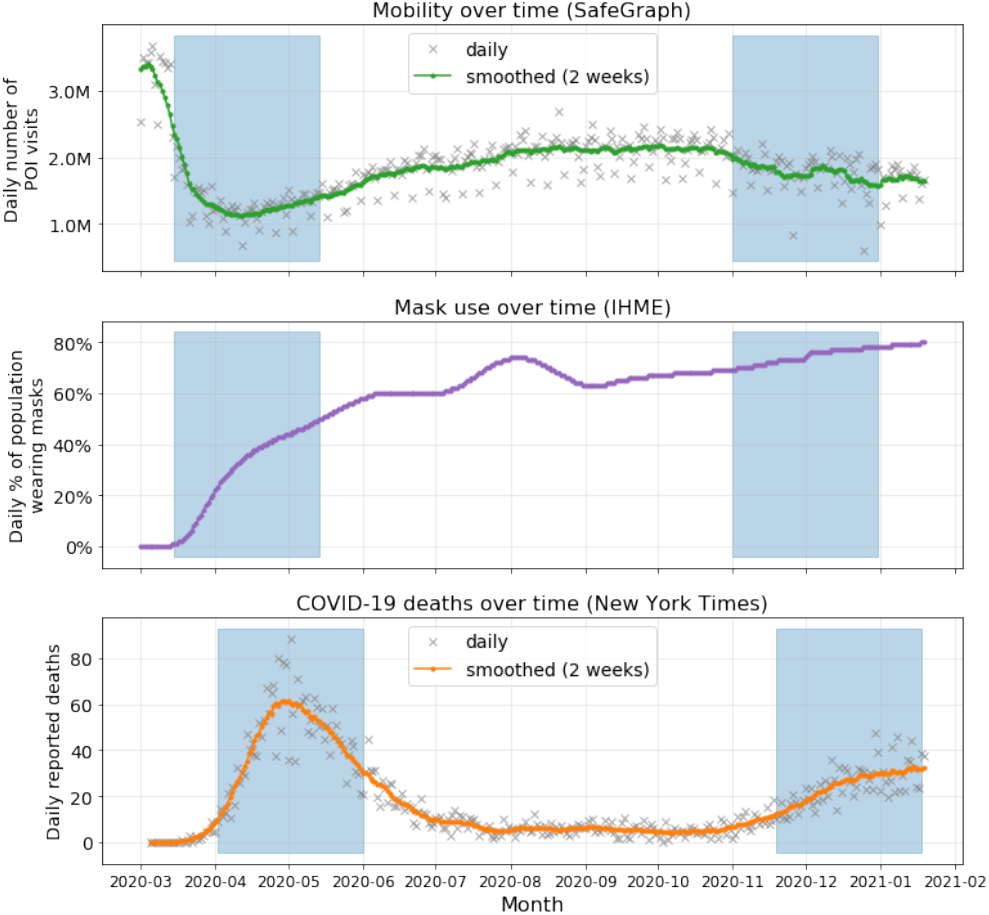
Three time-varying data sources – mobility, mask use, and daily COVID-19 deaths – shown for the Washington DC MSA. The highlighted regions indicate the periods that we test for model validation, one from the first wave and one from the second wave of infections (Section 3.1). There is an 18-day offset between the highlighted regions for input data (mobility, mask use) and deaths, due to the modeled delay between becoming infectious and date of death (Section 2.2).

In this work, we focus on three of the largest metropolitan statistical areas (MSAs) in Virginia: Washington-Arlington-Alexandria, DC-VA-MD-WV (hereby referred to as the “Washington DC” MSA), Virginia Beach-Norfolk-Newport News, VA-NC (“Eastern”), and Richmond, VA (“Richmond”). Across these MSAs, we model 63,744 POIs and 7,609 census block groups (CBGs) in total, with over 3 billion hourly edges between them (Table 1).

**Table 1:**
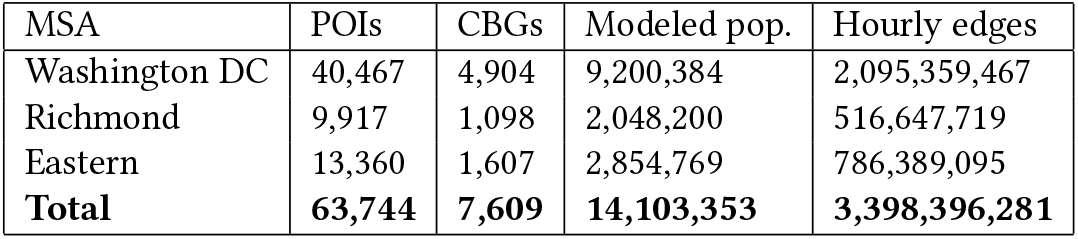
Summary of mobility networks. Modeled pop. indicates the total population living in the modeled CBGs. Hourly edges counts the number of non-zero edge weights in the mobility network from 12am November 1 to 11pm December 31, 2020 (the second wave period that we fit).

| MSA | POIs | CBGs | Modeled pop. | Hourly edges |
| --- | --- | --- | --- | --- |
| Washington DC | 40,467 | 4,904 | 9,200,384 | 2,095,359,467 |
| Richmond | 9,917 | 1,098 | 2,048,200 | 516,647,719 |
| Eastern | 13,360 | 1,607 | 2,854,769 | 786,389,095 |
| **Total** | **63,744** | **7,609** | **14,103,353** | **3,398,396,281** |

#### Mask-wearing data (IHME)

We use mask-wearing data from the Institute for Health Metrics and Evaluation (IHME) website,^5^ which provides daily estimates at the state level of the percentage of the population wearing masks. In Virginia, we see the most dramatic change in mask-wearing near the beginning of the pandemic, from 0% of the population wearing masks in mid-March to 60% by the end of May 2020 (Figure 2).

#### COVID-19 deaths (New York Times)

To calibrate our model, we compare its predicted death counts to data on reported deaths. We use *The New York Times’* COVID-19 dataset,^6^ which contains daily reported deaths per US county. For each MSA that we model, we sum over the county-level counts to produce overall counts for the entire MSA. As shown in Figure 2, Washington DC—along with much of the US—experienced two major waves of infections, one in the spring of 2020 and the second near the end of the year.

#### Demographic data (US Census)

We utilize data about each CBG from the American Community Survey (ACS) of the US Census Bureau. We use the 5-year ACS data (2013–2017) to extract the median household income, the proportion of white residents, and the proportion of Black residents of each CBG. For the total population of each CBG, we use the most-recent one-year estimates (2018); one-year estimates are noisier, but we wanted to minimize systematic downward bias (due to population growth) by making them as recent as possible. The model uses CBG populations as input, but it does not use income or race during the simulation. Instead, we use income and race data to analyze the model’s output; for example, to compare the predicted infection rates of lower-income and higher-income CBGs (Section 3.2).

### 2.2 Epidemiological model

#### Mobility network

We overlay a disease transmission model on a dynamic mobility network, which is represented as a complete undirected bipartite graph *𝒢* = (*𝒱, ϵ*) with time-varying edges. The nodes *𝒱* are partitioned into two disjoint sets *𝒸* = {*c*_1_, …, *c*_*m*_}, representing *m* CBGs, and 𝒫 = {*p*_1_, …, *p*_*n*_=}, representing *n* POIs.

The weight 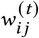 on an edge (*c*_*i*_, *p*_*j*_) indicates the number of people from CBG *c*_*i*_ who visited POI *p*_*j*_ in hour *t*; we refer the reader to Chang et al. [8] for the details of how we derive the hourly edge weights from SafeGraph data. From US Census data, each CBG *c*_*i*_ is labeled with its population 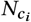 and from SafeGraph data, each POI *p*_*j*_ is labeled with its area 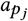 and median dwell time 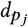

#### Model dynamics

We assume that every individual is in one of four disease states at any given time: susceptible (*S*), exposed (*E*), infectious (*I*), or removed (*R*). Susceptible individuals can acquire the virus through contact with infectious individuals. They then enter the exposed state, during which they have been infected but are not yet infectious. Individuals transition from exposed to infectious at a rate inversely proportional to the mean latency period. Finally, they transition from the infectious state to the removed state at a rate inversely proportional to the mean infectious period.In the removed state, they can no longer infect others or become infected again (e.g., because they have recovered or died).

In our model, each CBG maintains its own SEIR states, with 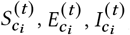 and 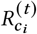 representing how many individuals in CBG *c*_*i*_ are in each disease state at hour *t*, and 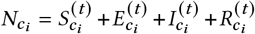 At each hour *t* we sample the transitions between states as follows:

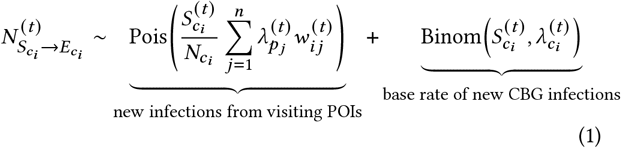

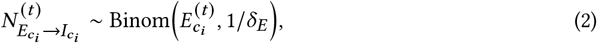

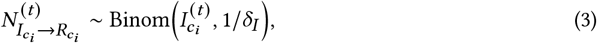

where 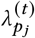 is the rate of infection at POI *p*_*j*_ at hour *t* 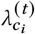 is the base rate of infection that is independent of visiting POIs; *δ*_*E*_ is the mean latency period; and *δ*_*E*_ is the mean infectious period.

#### The number of new exposures

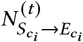 We assume that any susceptible visitor to POI *p*_*j*_ at hour *t* has the same independent probability 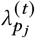 of being infected. Since there are 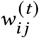 visitors from CBG *c*_*i*_ to POI *p*_*j*_at hour *t*, and we assume that a 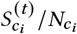 fraction of them are susceptible, the number of new infections among these visitors is distributed as 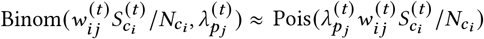. The number of new infections among all outgoing visitors from CBG *c*_*i*_ is therefore distributed as the sum of the above expression over all POIs, 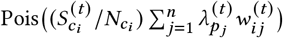.

We define the infection rate 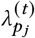 at POI *p*_*j*_ at hour *t* as the product of three time-varying factors: (1) the transmission rate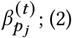 the density of infectious visitors 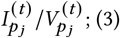 a mask-wearing factor (1-*ϵπ*^(*t*)^)^2^ [12], where *ϵ* ∈ [0, 1] represents mask efficacy and *π* ^(*t*)^ indicates the proportion of the MSA population wearing a mask at hour *t* (see Section A.1.2 for details).

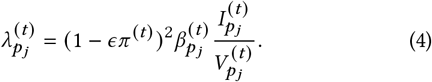

The transmission rate is 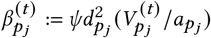, where *ψ* is a transmission constant (shared across all POIs) that we fit to data, the dwell time 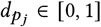 is the average fraction of an hour a visitor spends at *p*_*j*_, and 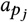 is the physical area of *p*_*j*_.

In addition to new infections from POIs, we model a CBG-specific base rate of new infections that is independent of POI visit activity This captures other sources of infections, e.g., household infections or infections at POIs that are absent from the SafeGraph data. At each hour *t*, every susceptible individual in CBG *c*_*i*_ has a base probability 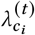 of becoming infected, where

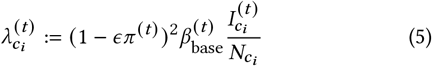

is the product of the mask-wearing scaling factor, the base transmission rate 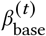, and the proportion of infectious individuals in *c*_*i*_ We parameterize *β*_base_ by defining a starting point 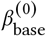 and ratio *r*_β_, so that *β*_base_ updates linearly from 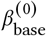 to 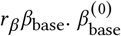 and *r*_β_ are free parameters, shared across all CBGs, which we fit to data We allow *β*_base_ to vary over time to capture changes in behavior outside of POIs (e.g., if home gatherings increase). However, we restrict the amount that it can vary by using a conservative range for *r*_*β*_, allowing *β*_base_ to change up to 30% (in either direction) over the 2-month periods that we simulate.

#### The number of reported deaths

We assume that a time-varying proportion of infections 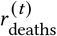 will result in reported deaths, and that they will be reported exactly δ_deaths_ = 432 hours (18 days) after the individual became infectious.^7^ From these assumptions, the predicted number of newly reported deaths from CBG *c*_*i*_> on day *d* is

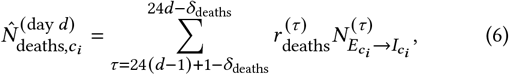

where we define 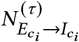 to be 0 when g *< <*1. This conversion from *SEIR* states to reported deaths allows us to fit our models on daily reported deaths, which we describe in the following section, as well as to initialize the *SEIR* states at the beginning of the simulation based on historical reported deaths (Section A.1.3).

The time-varying reported death rate 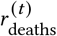 is the product of three factors: (1) the initial infection fatality rate IFR_0_ = 0.0068 [24]; (2) the relative reduction in the infection fatality rate 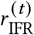 over time; (3) the proportion of COVID-19 deaths that are detected 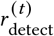 We use existing estimates of 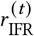, which, based on hospital fatality rates, estimate that the infection fatality rate was nearly halved by the detect summer of 2020 [15]. To estimate 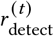, we compute the weekly ratio of reported COVID-19 deaths in the US, from *The New York Times*, over weekly select-cause excess deaths, as estimated by the National Center for Health Statistics (NCHS).^8^ The NCHS provides, for select causes of deaths determined to be related to COVID-19 (e.g., pneumonia, heart failure), the expected (based on 2015-2019) and actual numbers of deaths due to these causes for each week since the pandemic began; the weekly excess select-cause deaths are the difference between the expected and actual (or 0, if the expected exceeds the actual). If we assume that all select-cause excess deaths can be attributed to COVID-19, then we find that the death detection rate 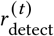 increased from around 20% at the beginning of the pandemic to 60%-80% after June 2020.

#### Model fitting

Most of our model parameters can either be estimated from data or taken from prior work (see Table A1 for a summary). This leaves 3 model parameters that we need to calibrate with data: the POI transmission constant, *ψ*, and the starting point and ratio for the base transmission rate, 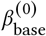 and *r*_*β*_. We calibrate these parameters per MSA by fitting to published numbers of confirmed deaths, as reported by *The New York Times* (NYT). NYT data provides the daily cumulative number of COVID-19 deaths per county, so first we sum over county-level counts to produce cumulative death counts for the MSA. Then, we convert this to the daily number of *new* deaths and apply two-week averaging to the raw daily counts to smooth over weekly effects (e.g., not reporting on weekends) and other sources of noise. We measure model fit based on new deaths per day (i.e., incident deaths), since model error and uncertainty can be severely underestimated when the fit is evaluated on cumulative curves [19].

Let 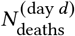 represent the (smoothed) number of reported deaths on day *d* in the MSA, and let 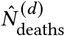 represent the model’s prediction for this quantity, which we compute by summing over the model’s CBG-level predictions (Equation 6). We compare our model predictions to the actual counts by computing the root-mean-squared-error (RMSE) between each of the days of our simulation:

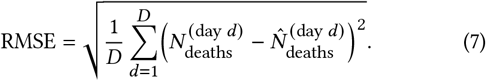

For each parameter set, we quantify model fit by running 30 stochastic realizations and averaging their RMSE.

Throughout this paper, we report aggregate predictions from the best-fitting parameter sets. For each MSA, we:

(1)Grid-search over 1,050 combinations of *ψ*, 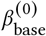, and *r*_*β*_

(2)Find the best-fit parameter set with the lowest average RMSE.

(3) Select all parameter sets that achieve an RMSE within 20% of the RMSE of the best-fit parameter set.

(4) Pool together all predictions across those parameter sets and all of their stochastic realizations, and report their mean and 2.5th/97.5th percentiles.

This procedure captures uncertainty from two sources: (1) stochastic variability between model runs with the same parameters, and (2) uncertainty in the model parameters. In Table 2, we describe the selected parameters for each of the MSAs that we model.

**Table 2:** Quantitative results from model fitting. For each model, we keep all parameter sets that achieve an RMSE within 20% of the RMSE of the best-fit parameter set. Here, we report the number of parameter sets kept per model, and the mean and range over the values for each parameter. nRMSE indicates the normalized RMSE, i.e., the RMSE divided by the MSA’s mean daily reported deaths over this period; we normalize in order to facilitate comparison across periods and MSAs. Note that the four rows in this table correspond to the four visualizations in the gure above.

| Period | MSA | # parameter sets | $\psi$ | $\beta_{\text{base}}^{(0)}$ | $r_\beta$ | nRMSE |
| --- | --- | --- | --- | --- | --- | --- |
| 1st wave | Washington DC | 3 | 4452.89 (3051.76-6204.30) | 0.013 (0.009-0.016) | 0.733 (0.700-0.800) | 0.053 |
| 2nd wave | Washington DC | 3 | 4817.52 (628.32-9006.72) | 0.027 (0.015-0.039) | 1.133 (1.100-1.200) | 0.072 |
| 2nd wave | Richmond | 5 | 4817.52 (628.32-9006.72) | 0.023 (0.009-0.039) | 1.100 (1.000-1.200) | 0.115 |
| 2nd wave | Virginia Beach | 13 | 7932.57 (628.32-11799.52) | 0.017 (0.003-0.045) | 0.962 (0.700-1.300) | 0.173 |

### 2.3 Large-scale implementation

In this section, we describe computational challenges that arise from implementing this system at scale, and discuss how we addressed them with engineering solutions.

#### Handling large mobility networks

The hourly POI-CBG networks store large amounts of data; for example, across the three Virginia MSAs we focus on in this work, their networks contain 3.4 billion hourly edges from November to December 2020 (Table 1). In order to reduce computation time, we run our network inference algorithm ahead of any disease modeling, and save the inferred edge weights so that they can be loaded later on. For each network, we save the weights separately per hour (as sparse matrices), so that the model only needs to load as many hours of data as necessary for the current simulation. The model dynamics bring their own challenges: in every hour, we need to estimate the infection rate per POI (Equation 4), and the base infection rate per CBG (Equation 5). However, because each POI’s hourly infection rate is conditionally independent of the other POIs’ infection rates (given the current *SEIR* states for each CBG), we can parallelize the hourly computations across POIs; for similar reasons, we can parallelize across CBGs and random seeds (i.e., stochastic realizations). These strategies greatly reduce simulation time; for instance, bringing the average runtime for a 2-month multi-seed simulation with Washington DC down to 5.5 minutes, even as each simulation requires computing 1.78 billion hourly, seed-specific POI infection rates and 215 million hourly, seed-specific CBG infection rates.

#### Scaling model experiments

One of the strengths of our approach is flexibility: for example, our dashboard allows policymakers to test any combination of opening 5 different POI categories to 4 different levels of mobility (Section 3.3). However, this flexibility also creates an exponential number of scenarios to simulate (4^5^ = 1, 024). Furthermore, for each scenario, we run 30 stochastic realizations for every parameter set; thus, with 9 parameter sets (Table 2), we need to run nearly 300,000 model realizations. As described above, part of the solution lies in our model implementation, which runs the stochastic realizations per parameter set in parallel. However, the key to completing these experiments is that we distribute the work across multiple computers, with collectively 288 cores. This allows us to run hundreds of simulations in parallel, compressing 2 years of compute time into a few days.

## 3 RESULTS

### 3.1 Model validation

First, we calibrated models for each of the three Virginia MSAs using input data from November 1 to December 31, 2020 and reported deaths from November 19 to January 18, 2021 (there is a 18-day offset between these ranges due to the lag from becoming infectious to date of death, δ_*c*_). In Figure 3b–d, we show that our models are able to accurately fit daily deaths for these MSAs during this time period. The fit is especially good for Washington DC, with a normalized RMSE of 7.2% (Table 2). The normalized RMSEs for the Richmond and Eastern MSAs are slightly higher at 11.5% and 17.3%, respectively; these regions were more challenging to fit during this time period due to the large amount of noise relative to their single-digit reported daily deaths (Figure 3d).

**Figure 3.**
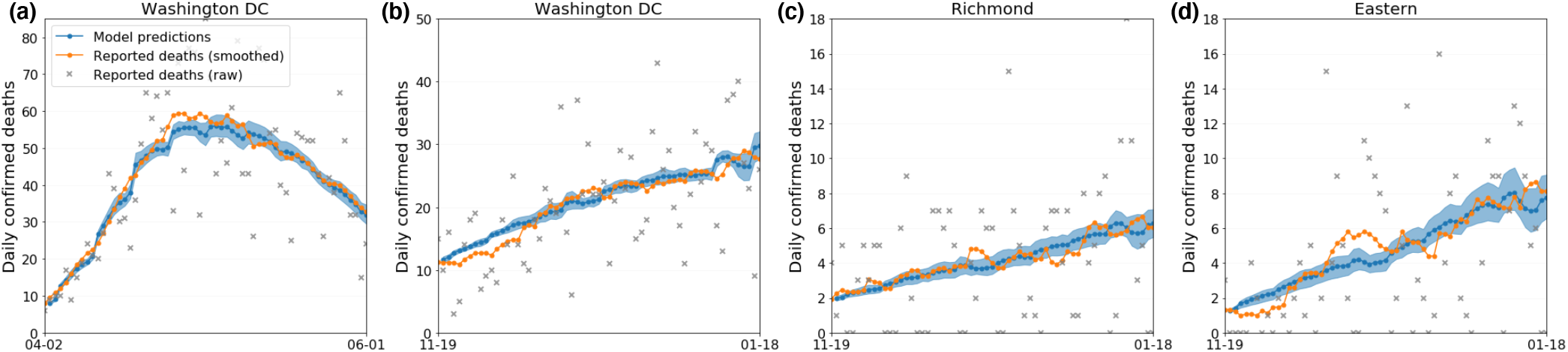
Model fit with “full” (non-ablation) model. We calibrate models for each of the three MSAs for the second wave period (b–d) and a model for Washington DC in the first wave (a). We apply 2-week averaging to the raw counts for daily reported deaths (grey) to smooth over weekly effects and noise. We fit the model to the smoothed daily deaths (orange). The shaded regions indicate the 2.5th to 97.5th percentiles over model parameters and stochastic realizations.

To further test our model, we conduct a series of extended analyses with Washington DC as our case study. We focus on Washington DC because its daily death counts are less noisy than the counts for the other two MSAs, due to its substantially larger size (it is 4 their sizes, and in fact the 6th largest MSA in the US). In addition to the November to January period that we fitted above, we fit our model using input data from March 15 to May 14, 2020 and reported deaths from April 2 to June 1, 2020.^9^ We choose these two periods since they overlap with the first and second waves of infection, and because they reflect vastly different points in the pandemic, with different distancing behaviors, proportions of people infected, weather conditions, and so on. For each time period, we also conduct ablation studies to test the importance of several model features. In the first ablation, we remove mobility data by fixing ψ, the POI transmission constant, to 0, and allow the model to search over a wider and finer grid for the base transmission rate,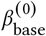. In the second ablation, we remove mask-wearing data by xing the mask-wearing proportion π to 0, but we search over the same grids as in the original model since π was not a free parameter. The third ablation keeps *β*_base_ constant for the entire time period, instead of allowing it to vary slightly over time. To do this, we fix *r*_*β*_ to 1, and search over a wider and finer grid for 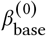

First, we find that our model can also accurately fit the non-linear daily deaths curve from the first wave (Figure 3a), with a normalized RMSE of 5% (Table 2). Furthermore, the model outperforms its ablations in both periods, although the impact of removing any feature is substantially larger in the first wave than the second, due to the larger changes in behavior early on. For example, removing mask-wearing data results in a severe increase in the model’s RMSE (+591%) during the first wave, where the increase in mask-wearing allowed the model to capture the downward trend in deaths in May 2020, even as mobility began to slightly climb during this period (Figure 2). Removing mobility during this time period also has a large effect, nearly doubling the model’s RMSE, and fixing *β*_base_ over time results in a 74% increase. In the second wave, the impacts are more subtle: removing mobility data, removing mask-wearing data, and fixing *β*_base_ over time result in 14%, 13%, and 5% increases in the model’s RMSE, respectively (Table A2).

### 3.2 Use cases

Our fitted model can be applied to a wide variety of use cases, including retrospective analyses investigating who was infected where and when, and forward-facing experiments that modify the model’s inputs to test hypothetical changes in policy or behavior in the near future. In this section, we provide a few examples that demonstrate the retrospective and forward-facing capabilities of our model, and highlight the usefulness of our fine-grained approach in capturing heterogeneity in risk across POIs and CBGs.

#### Analyzing disparities in infection rates

Our first use case is an example of retrospective analysis. After fitting the model, we might be interested in studying what the model learned about the infection rates for lower-income versus higher-income CBGs, since it is well-reported in the real world that lower-income neighborhoods have been impacted more severely by COVID-19 [34]. To analyze this, we stratify CBGs by median household income and compare the cumulative infection rates over time (anyone in *E, I*, or *R*) of the CBGs in the bottom income decile versus top income decile. We find that the model correctly predicts a large disparity between the bottom and top income deciles in Washington DC [30] (Figure 4). This gap is especially striking in the first wave period: from March 15 to May 14, cumulative predicted infections per 100,000 increased around 60% more for the bottom income decile than the top income decile (9,900 versus 6000). This difference can only be attributed to differing mobility patterns, since the CBGs were initialized to very similar levels of infection at the beginning of the simulation and all other exogenous model parameters are shared across CBGs. This matches our first wave findings from the original work [8], where we found across 10 of the largest MSAs in the US (including Washington DC), CBGs in the bottom income decile always had a higher infection rate by the end of the simulation than the top income decile. What we found in the mobility data that explained this difference was that lower-income CBGs could not reduce their mobility as much and, even within the same POI category, the POIs that they visited tended to be more crowded and thus higher-risk. In the second period, the bottom income decile begins at a disadvantage, reflecting actual cumulative deaths by the beginning of November 2020 (see Section A.1.3 for how we initialized *SEIR* states using historical reported deaths). However, the disparity continues to grow: by the end of December 2020, predicted cumulative infections per 100,000 increased 30% more for the bottom income decile than the top income decile (8,900 versus 6,800). This dffierence can be partially attributed to mobility patterns, but also to the self-compounding nature of the virus. Since lower-income CBGs begin with a larger number of infectious individuals, those individuals will generate more infectious people, exacerbating the disparity. Our model can capture these self-compounding dynamics, as well as demonstrate how mobility contributes to—but could also be used to mitigate—these health disparities.

**Figure 4.**
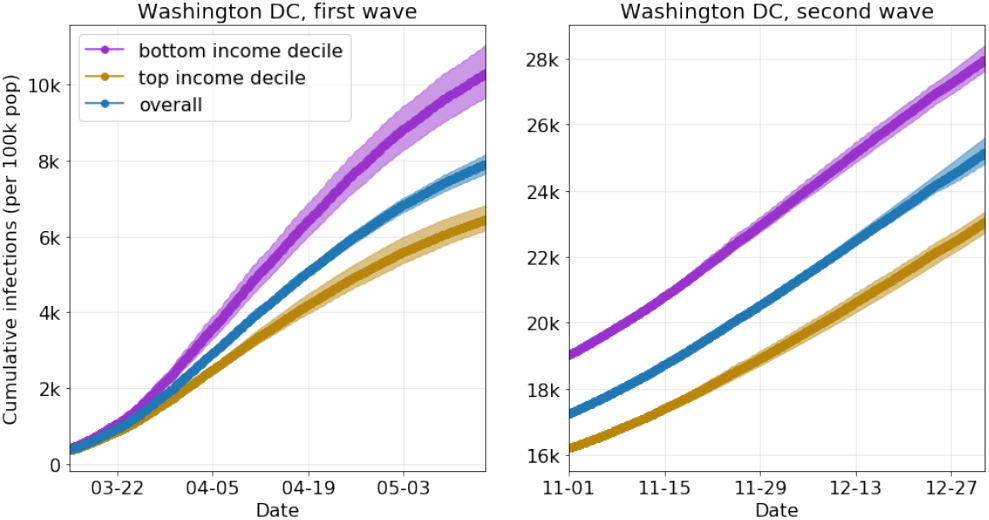
Retrospective analysis of disparities in infection rates, comparing Washington DC CBGs in the bottom decile (purple) vs. top decile (gold) of median household income. The overall population (blue) is also shown. The shaded regions indicate the 2.5th to 97.5th percentiles over model parameters and stochastic realizations.

#### Opening POI categories to different degrees

Our second use case is an example of a forward-facing experiment. One common strategy by policymakers has been to implement varying restrictions on different business sectors [7], so here we explore the effects of opening POI categories to differing levels of mobility. To test this, we run a simulation starting from November 1, 2020, using the models that we already calibrated (Section 3.1). Then, we modify the mobility networks starting on January 1, 2021, and run the model forward with the modified mobility network for four weeks. During those four weeks, each POI category either maintains its current levels of mobility, or we specify it to have a certain fraction of its “normal” mobility levels, based on SafeGraph data from January 2019. We perform this experiment with 5 POI categories: (1) Restaurants; (2) Essential Retail (grocery stores, convenience stores, drug stores); (3) Gyms; (4) Religious Organizations; and (5) Retail (clothing stores, hardware stores, book stores, pet stores, etc.). For each category, we consider 4 possible mobility settings: maintaining the current mobility level, or keeping 0%, 50%, or 100% of 2019 mobility. In order to provide a wide array of options to policymakers, we test every combination of POI category and mobility setting (1,024 options per MSA).

These experiments allow us to quantify the trade-of between visits and infections. For example, if every POI continued at current levels of mobility, our model predicts that the Washington DC MSA would experience around 267,000 new infections (2.9% of the population) in January 2021. If, instead, the Restaurant POIs returned to 2019 levels of mobility starting on January 1, we would see a 34% increase in POI visits, but also a 179% increase in predicted new infections. In contrast, if the Essential Retail POIs returned to 2019 levels of mobility, we would only see a 4% increase in POI visits and a 8% increase in predicted new infections. These differences are partially because there are far more Restaurant than Essential Retail POIs in the Washington DC MSA (10,545 versus 1,606), but also because Restaurant POIs saw a larger drop in mobility during the pandemic, so returning them to “normal” levels of mobility would have a larger impact. Testing every combination of category and mobility also allows us to inspect interactions between categories. For example, if we returned Restaurant *and* Essential Retail POIs to 2019 mobility levels, we see a 198% increase in predicted new infections; this is higher than the sum of the predicted infections from opening each one on its own (which would be 187% = 179% + 8%). This result highlights why we simulate each combination of mobility levels instead of adding individual impacts, since the whole impact of a policy can be greater than the sum of its parts.

### 3.3 Dashboard

Our dashboard provides an interface to our model results which public health officials can use to assess the impact of mobility on COVID-19 transmission. We designed our dashboard through iterative meetings with VDH, where they provided valuable feedback on how the interface could be made more intuitive and which visualizations would be most effective in conveying the impact of changing mobility levels. The resulting layout is divided into five parts, as shown in Figure 5; we discuss each part in detail below.

**Figure 5.**
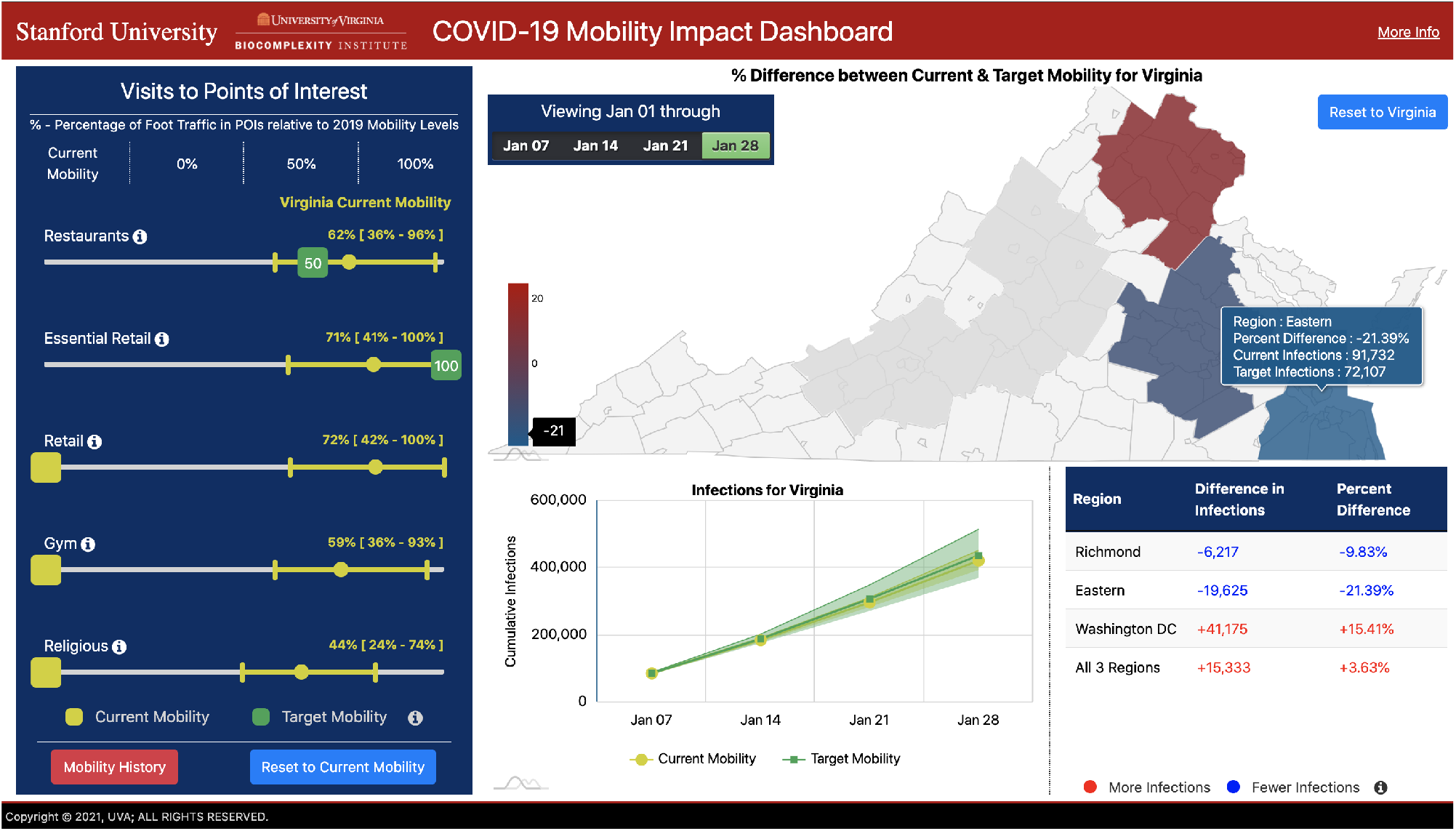
Our dashboard is divided into 5 parts: the POI Navigation Bar (left); the Map Panel (upper right); the Chart Panel (lower center); the Data Panel (lower right). and the Mobility History Panel (popup)

#### Visits to Points of Interest Navigation Bar

The POI Navigation Bar is the control center of the tool. From here, users can either view current mobility levels by POI category or use sliders to set “target” mobility levels to 0%, 50% or 100%, indicating the fraction of 2019 mobility levels to use. For each category, we have a current mobility ratio associated with each POI (i.e., how their current mobility compares to their mobility levels from 2019); to communicate heterogeneity in current mobility ratios across POIs within each category, we show each category’s median and interquartile range of current mobility ratios as yellow markers on the slider axes. When the application loads, the selected region defaults to Virginia, and the current mobility levels displayed are the average of the mobility levels for the three MSAs.

#### Map Panel

As the user changes target mobility on the POI Navigation Bar, the colors of each MSA region on the map also change to reflect increases (more red) or decreases (more blue) in the number of predicted infections. This provides a visual indicator of where a mobility change is likely to have the greatest impact; to get more precise measurements, the user can hover the mouse over each MSA to display predicted infections at current and target mobility levels, as well as the percent difference between them. Model predictions can be viewed at 1, 2, 3, or 4 weeks following the “intervention date” (in this case, January 1, 2021). We provide this option because the impact of the change in mobility on predicted infections can evolve over time, typically accumulating in magnitude. For example, relative to maintaining current levels of mobility, setting Restaurant POIs in Washington DC to 100% of 2019 mobility levels results in a 76% increase in predicted infections after 2 weeks, a 136% increase after 3 weeks, and, as mentioned in the previous section, a 179% increase after 4 weeks. The user can also click on a specific MSA to select it; this updates the POI Navigation Bar with current mobility levels for the selected MSA, and the Chart Panel with predictions associated with the selection. The user can click on the “Reset to Virginia” button to return to the three MSAs as a collective unit.

#### Chart Panel

While the Map Panel displays the cumulative impact of the target mobility change for one weekly period at a time, the Chart Panel displays the time series for cumulative predicted infections at current and target mobility levels across the full four-week period along with their 95% confidence intervals over model parameters and stochastic realizations. This makes it easier to visualize how much predicted infections at target mobility are expected to deviate from predictions at the current mobility level.

#### Data Table Panel

The Data Table shows the difference between predicted infections given current and target mobility levels. It is responsive to the selection of different target mobility levels on the POI Navigation Bar, as well as the selected week on the Map Panel. This feature allows the user to conveniently assess the quantitative impact of changing mobility levels, both at the MSA and overall Virginia levels.

#### Mobility History

The Mobility History Panel is revealed upon selection of the “Mobility History” button on the POI Navigation Bar. This report provides a history from January 2019 to present day of weekly POI visits, aggregated by MSA and POI categories, which helps policy health experts better contextualize the target mobility levels in the POI Navigation Bar. We provide a screenshot from and additional details about this panel in Section A.2.

## 4 RELATED WORK

### Mobility and COVID-19 modeling

The COVID-19 pandemic and its corresponding social distancing measures have drastically affected human mobility patterns [13, 16]. To accurately capture the dynamics of COVID-19 transmission and infection, epidemiological models must account for these changes in mobility. Many such models have been proposed in the last year: for example, it is common for models to use some aggregate measure of real-time mobility to modulate transmission rates [9, 17, 20]. Others have focused on using historical data to model the initial spread of the disease before social distancing measures were put into place [21, 29], or on using synthetic data for analytical purposes [5, 18]. In this work, we extend the model from Chang et al. [8], as it uses mobility data that is both fine-grained (e.g., at the individual POI and CBG level) and up-to-date, which enables us to predict the effects of different mobility restriction measures at the level of granularity required for policy-making [4, 6].

### COVID-19 tools for policymakers

Recently, there has been an intense interest in developing easy-to-use interfaces to support computational infectious diseases epidemiology. Much of the effort is focused on providing surveillance information [11, 28]; for example, several apps have been built to monitor trends in cases over time [35], tracking their growth rate [1] or aiming to detect clusters [14]. Other modeling tools that are being used for the COVID-19 pandemic include EpiC and Gleamviz [32], for global mobility and epidemic simulations; DiCon, for optimization and control problems related to epidemic dynamics [25]; and, most related to our tool, FRED [26] and GAIA [27], which are open source systems that support research in networked epidemiology. Our team has also been developing epidemiological applications in support of policy-makers for over 15 years, including SIBEL, an epidemic modeling tool that allows users to experiment with fine-grained interventions [2, 3, 10, 22, 23], and EpiViewer, a tool for visualizing epidemic time series [31]. What sets the tool described in this paper apart from the others is the way it incorporates fine-grained mobility data with the SEIR model, enabling a more detailed ability to create and test interventions. Our tool also focuses on near real-time response; this marks a crucial evolution from earlier efforts that were largely used for planning studies.

## 5 CONCLUSION

We have introduced a decision-support tool that allows policy-makers to inspect the predicted impacts of thousands of different policies, specific to their jurisdictions. Our tool utilizes large-scale data and epidemiological modeling to simulate the effects of negrained changes in mobility on infection rates, and leverages a robust computational infrastructure to run model experiments at scale. As policymakers face difficult challenges ahead, our tool will provide them with much-needed analytical machinery to assess tradeoffs between future infections and mobility restrictions.

Our approach is not without its limitations, which we have discussed with policymakers. For instance, our mobility data from SafeGraph does not cover all POIs or populations (e.g., children), and our model makes necessary but simplifying assumptions about the dynamics of disease transmission. Furthermore, we specialize in modeling the effects of changes in mobility on infection rates, but not all of the mobility policies that we analyze are directly actionable; for example, changing current mobility levels to 50% of 2019 mobility. Further work is required to analyze how to lever tools in policymakers’ toolbox to actually reach target levels of mobility. Despite these limitations, our approach captures a valuable piece of the puzzle, as we provide policymakers with a quantitative and comprehensive near real-time analysis of the effects of mobility on transmission. As we move forward, we will build dashboards for other US states and regions, and continue developing new use cases and technical advances for our model so that we can best support the needs of policymakers around the country.

## Data Availability

Mobile phone mobility data are freely available to researchers, non-profit organizations and governments through the SafeGraph COVID-19 Data Consortium. All other data come from publicly available sources; we provide links to these data below. The dataset from The New York Times consists of aggregated COVID-19-confirmed case and death counts collected by journalists from public news conferences and public data releases. We also use data from the National Center for Health Statistics (NCHS) to estimate national excess deaths. Our state-level mask-wearing data comes from the Institute for Health Metrics and Evaluation's (IHME) public dashboard. Additional data about US census block groups come from the US American Census Survey. Our code is also available online at https://github.com/snap-stanford/covid-mobility-tool.

https://www.safegraph.com/covid-19-data-consortium

https://github.com/nytimes/covid-19-data

https://www.cdc.gov/nchs/nvss/vsrr/covid19/excess_deaths.htm

https://covid19.healthdata.org/united-states-of-america

https://www.census.gov/programs-surveys/acs

## ACKNOWLEDGEMENTS

The authors would like to thank members of the Biocomplexity COVID-19 Response Team and the Network Systems Science and Advanced Computing (NSSAC) Division and members of the Biocomplexity Institute and Initiative, University of Virginia, for useful discussion and suggestions. This work was partially supported by NSF BIG DATA Grant IIS-1633028, NSF Grant No. OAC-1916805, NSF Expeditions in Computing Grant CCF-1918656, CCF-1917819, NSF RAPID CNS-2028004, NSF RAPID OAC-2027541, NSF OAC-1835598 (CINES), NSF OAC-1934578 (HDR), NSF CCF-1918940 (Expeditions), NSF IIS-2030477 (RAPID), Stanford Data Science Initiative, Wu Tsai Neurosciences Institute, Chan Zuckerberg Biohub, United Health Group, US Centers for Disease Control and Prevention 75D30119C05935, University of Virginia Strategic Investment Fund award number SIF160, and Defense Threat Reduction Agency (DTRA) under Contract No. HDTRA1-19-D-0007. S.C. was supported by an NSF Graduate Fellowship. P.W.K. was supported by the Facebook Fellowship Program. J.L. is a Chan Zuckerberg Biohub investigator. Any opinions, findings, and conclusions or recommendations expressed in this material are those of the author(s) and do not necessarily reflect the views of the funding agencies.

## A APPENDIX

### A.1 Details to our approach

#### A.1.1 POI and CBG inclusion criteria

Since we model one MSA at a time, we need to dene ltering criteria to determine which POIs and CBGs to include in the model. Fifirst, we include all POIs that meet the following requirements: (1) the POI is located in the MSA; (2) SafeGraph has visit data for this POI for every hour from 12am on September 1 2020 to 11pm on November 30 2020^10^; (3) SafeGraph has recorded this POI’s median dwell time and visitors’ home CBGs for at least one week from January to October 2020; (4) SafeGraph provides this POI’s area in square feet; (5) this POI is not a “parent”^11^ POI. After determining the set of included POIs, we keep the union of CBGs that are located in the MSA and those that had at least one recorded visit to at least 100 of the kept POIs; this means that CBGs from outside the MSA may be included if they visit this MSA frequently enough.

#### A.1.2 Derivation for mask-wearing

Here, we explain how we incorporated mask-wearing into our model dynamics. Fifirst, note that without mask-wearing, both infection rate equations (4 and 5) can be written in the generalized form *βI*/*N*, where *β* is the transmission rate, *I* is the number of infectious individuals present, and *N* is the total number of individuals present. Furthermore, the expected number of new infections is *S*(*βI*/*N)*, where (*S* is the number of susceptible individuals present. Following Eikenberry et al. [12], let *π* ∈[0, 1] represent the fraction of the population wearing a mask. We can divide the infectious and susceptible populations into the infectious-unmasked group, *I*_*U*_= (1-π)*I*; the infectious-masked group, *I*_*M*_ := *πI*; the susceptible-unmasked group, *S*_*U*_= (1-*π*)*S;* and the susceptible-masked group *S*_*M*_*=πS*.Let the random vari able *X*_*U*_ represent the number of unmasked susceptible visitors who become infected, and let *X*_*M*_; represent the number of masked susceptible visitors who become infected. Then, we can derive the expected values of these variables by separating the cases where they become infected by an unmasked infectious person versus by a masked infectious person:

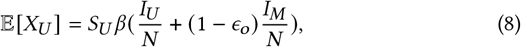

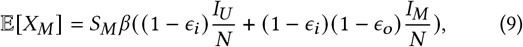

where *ϵ*_o_ ∈[0, 1] is the “outward” eciency of the mask (how much it prevents a masked infectious person from transmitting) and *ϵ*_*i*_ ∈[0, 1] is the “inward” effciency (how much the mask protects a masked susceptible person from catching the disease). If we assume *ϵ*_*o*_; *= ϵ*_*i*_ *= ϵ*, as [12] do in their experiments, and substitute back in the definitions of *I*_*U*_, *I*_*M*_; *S*_*U*_ *S*_*M*_, we find that the total expected number of infections *X* simplifies to

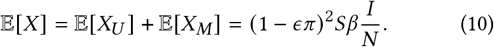

In other words, we simply scale the original expected number of new infections by a factor of (1 - *ϵ π)* ^2^. Plugging this general form back into the POI and base infection rates yields equations 4 and 5.

#### A.1.3 Model initialization

We use the county-level death counts from *The New York Times* to initialize the *SEIR*; states at the beginning of our simulations. For each county in the MSA, first we convert its cumulative death counts to daily new deaths, and apply 2-week smoothing to the raw counts (as we did to the MSA-level counts). For a county *Y*, let 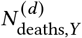 represent its smoothed actual number of new deaths on day 3. Our goal is to use this timeseries to estimate 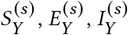, and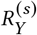, the number of people in the county in eac.h dise.ase .state for so.me simulation start hour *s*

Fifirst, recall that our model assumes deaths are reported exactly *δ*_deaths_/24 = 18 days after the person becomes infectious, and that on day *d* (*s*) = ⌊*s*/24⌋, a fraction 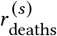 of the newly infectious cases will eventually result in reported deaths. If we assume 1 / 24 of the infections on day *d* (*s*)occurred at hour *s*, then the number of individuals in county. who became newly infectious at hour *s* must 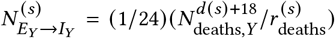. Furthermore, our model assumes that exposed individuals always have a 1 / *δ*_*E*_ probability of transitioning into the infectious state, so the maximum likelihood estimate of 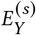 is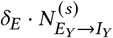. Since it takes on average *δ*_*E*_ hours for individuals to transition from exposed to infectious, then we estimate 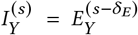 Similarly, since it takes on average *δ*_*I*_ hours for p.eople to. transition from infectious to removed, we set 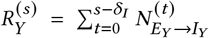, where *t* = 0 represents the start of the pandemic. In other words, by hour *s*, we assume everyone who transitioned into *I* before hour *s-δ*_*I*_ has reached *R*. Finally, we 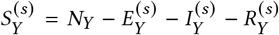. We note that these are rough estimates, but the uncertainty captured by our parameter selection and stochastic realizations should more than cover the uncertainty carried in our initialization procedure. Furthermore, the estimates produced by this method align with what we would expect: for example, it predicts that by November 1, 2020, across all counties in the US, the median county-level proportion of the population in *R* was 19%, with an interquartile range of 9%–34%.

**Table A1:** Model parameters. If the parameter has a xed value, we specify it under Value; otherwise, we write “Variable” to indicate that it varies across CBG / POI / MSA. Eikenberry et al. [12] estimate that inward mask efficiency could range from 20-80% for cloth masks, and outward efficiency could range from 0-80%, with 50% perhaps typical; thus, we set n to 0.5.

| Parameter | Description | Value (Source) |
| --- | --- | --- |
| $\delta_E$ | mean latency period | 96 hours [21] |
| $\delta_I$ | mean infectious period | 84 hours [21] |
| $\delta_{\text{deaths}}$ | lag from becoming infectious to date of death | 432 hours (18 days) [33] |
| $r_{\text{deaths}}$ | fraction of deaths reported at time $t$ | Variable (NCHS, NYT) |
| $\beta_{\text{base}}^{(0)}$ | starting point for base transmission rate, $\beta_{\text{base}}$ | Variable (Estimated) |
| $r_\beta$ | ratio controlling how much $\beta_{\text{base}}$ will vary | Variable (Estimated) |
| $N_{c_i}$ | population size of CBG $c_i$ | Variable (US Census) |
| $\psi$ | scaling factor for POI transmission | Variable (Estimated) |
| $a_{p_j}$ | area of POI $p_j$ in square feet | Variable (SafeGraph) |
| $w_{ij}^{(t)}$ | # visitors from CBG $c_i$ to POI $p_j$ at time $t$ | Variable (SafeGraph) |
| $\pi^{(t)}$ | fraction of population wearing a mask at time $t$ | Variable (IHME) |
| $\epsilon$ | mask efficiency | 0.5 [12] |

**Table A2:** Results from ablation studies. We tested the Washington DC model in two time periods, comparing the full model versus its ablations (Section 3). We find that the full model achieves lower RMSEs than each of the ablations in both time periods, and the impact of removing any feature is substantially larger when fitting the first wave.

| Model | 1st wave, RMSE | 2nd wave, RMSE |
| --- | --- | --- |
| full | 2.238 | 1.486 |
| no mobility | 4.361 | 1.796 |
| no mask | 15.473 | 1.777 |
| fixed $\beta_{\text{base}}$ | 3.895 | 1.600 |

For a CBG *c*_*i*_ in county *y*, we set its initial states to match the proportions of the county’s states; for example,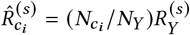. However, due to uncertainty in reported deaths and in our estimation method, after setting initial estimates for all CBGs in the MSA, we shrink each CBG’s estimate for every disease state toward the mean over all CBGs. For example, ultimately we set

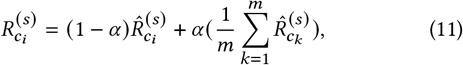

where *m* is the total number of CBGs in the MSA and *α* ∈ [0, 1] is a shrinkage parameter that controls how much we shrink toward the mean. Shrinkage allows us to be more conservative about our estimates, especially for CBGs with unusually high or low estimates for any of the disease states. Since we have greater uncertainty in reported deaths early in the pandemic, for the first wave period that we model (March to May 2020), we set *α* = 0.5, and for second wave period (November 2020 to January 2021), we set *α* = 0.1.

**Figure A1:**
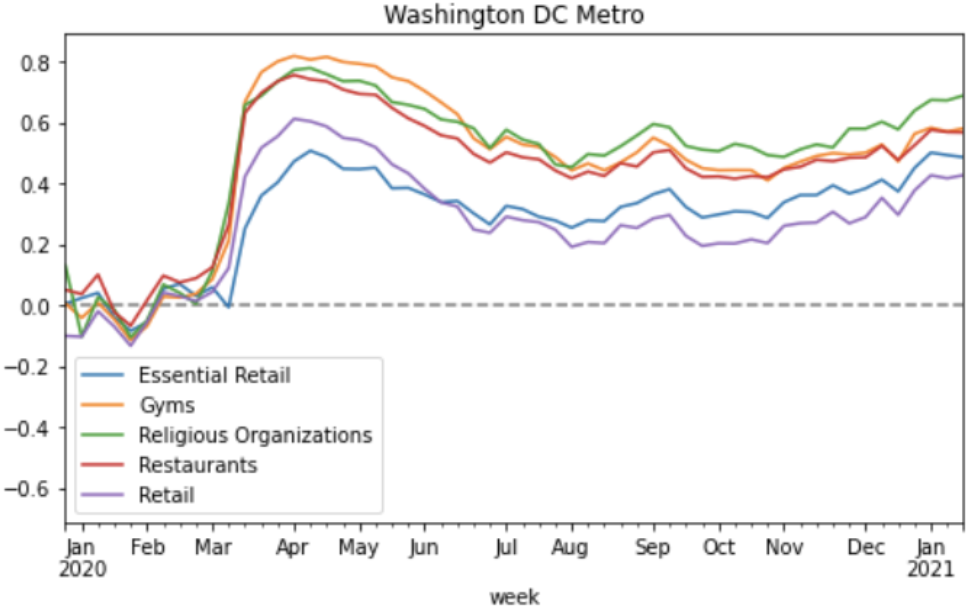
This plot showing the relative change in foot traffic per category over time is an example of the subreports available from the dashboard’s Mobility History Panel.

### A.2 Dashboard’s Mobility History Panel

The Mobility History report is available upon selection of the “Mobility History” button on the POI Navigation Bar. It provides aggregated POI visits per MSA and POI category, based on SafeGraph data, for every week from 2019 through the present time. In addition to providing raw data in tabular format, the report includes different visualizations of that data, including graphs of mobility counts in 2019 and 2020; the proportion of 2020-2021 foot traffic vs. 2019-2020 foot traffic by POI category, including bar graphs for the most recent week in the set for easier visualization; and the percent difference by POI category (Figure A1). This allows public health experts to review mobility trends and compare them to other COVID-19 indicators to make correlations and help inform them as they plan their guidance.

## Footnotes

1 Our code is available at https://github.com/snap-stanford/covid-mobility-tool.

2 https://docs.safegraph.com/v4.0/docs/places-schema.

3 https://docs.safegraph.com/v4.0/docs/weekly-patterns.

4 https://docs.safegraph.com/v4.0/docs/social-distancing-metrics.

5 https://covid19.healthdata.org/united-states-of-america/virginia?view=mask-use.

6 https://github.com/nytimes/covid-19-data.

7 It is simplifying to assume that the delay is fixed, but we performed sensitivity analyses in the original work that showed that model predictions would remain essentially identical if delays were sampled stochastically from a distribution.

8 https://www.cdc.gov/nchs/nvss/vsrr/covid19/excess_deaths.htm.

9 We choose two contrasting periods to t, instead of fitting the entire time period from March 2020 to January 2021, because fitting the entire time period would have required loading over 8,000 hourly mobility network weights, which would be around 100GB.

10 We use this range because we want to ensure that the POI was recently active, and we use this range even as we model periods from both the first and second wave of. infections because we want to keep the set of POIs xed across experiments.

11 Parent POIs consist of a small fraction of POIs that overlap with and include visits from their “children” POIs (for example, malls). To avoid double-counting visits, we. remove all parent POIs from the dataset.

## REFERENCES

[1] S. Barone et al. 2020. Building a statistical surveillance dashboard for COVID-19 infection worldwide. Quality Engineering (2020).

[2] C. Barrett et al. 2007. A scalable data management tool to support epidemiological modeling of large urban regions. In International Conference on Theory and Practice of Digital Libraries. Springer, 546–548.

[3] R. Beckman et al. 2014. Isis: A networked-epidemiology based pervasive web app for infectious disease pandemic planning and response. In KDD ‘14. 1847–1856.

[4] S. G. Benzell, A. Collis, and C. Nicolaides. 2020. Rationing social contact during the COVID-19 pandemic: Transmission risk and social benefits of US locations. PNAS (2020).

[5] P. Block et al. 2020. Social network-based distancing strategies to flatten the COVID-19 curve in a post-lockdown world. Nature Human Behaviour (2020).

[6] C. O. Buckee et al. 2020. Aggregated mobility data could help ght COVID-19. Science 368, 6487 (2020), 145.

[7] D. Bunis and J. Rough. 2021. List of Coronavirus-Related Restrictions in Every State. AARP: Politics & Society (2021).

[8] S. Chang, E. Pierson, P.W. Koh, et al. 2020. Mobility network models of COVID-19 explain inequities and inform reopening. Nature 589, 82–87 (2020).

[9] M. Chinazzi et al. 2020. The effect of travel restrictions on the spread of the 2019 novel coronavirus (COVID-19) outbreak. Science 368, 6489 (2020), 395–400.

[10] S. Deodhar et al. 2014. An interactive, Web-based high performance modeling environment for computational epidemiology. TMIS 5, 2 (2014), 1–27.

[11] E. Dong, H. Du, and L. Gardner. 2020. An interactive web-based dashboard to track COVID-19 in real time. The Lancet infectious diseases 20, 5 (2020), 533–534.

[12] S. Eikenberry et al. 2020. To mask or not to mask: Modeling the potential for face mask use by the general public to curtail the COVID-19 pandemic. Infectious Disease Modeling 5 (2020), 293–308.

[13] S. Gao, J. Rao, Y. Kang, Y. Liang, and J. Kruse. 2020. Mapping county-level mobility pattern changes in the United States in response to COVID-19. SIGSPATIAL Special 12, 1 (2020), 16–26.

[14] A. Hohl et al. 2020. Daily Surveillance of COVID-19 using the Prospective Space-Time Scan Statistic in the United States. Spatial and Spatio-temporal Epidemiology 34 (06 2020), 100354.

[15] T. Holden, R. Richardson, P. Arevalo, W. Duffus, M. Runge, E. Whitney, L. Wise, N. Ezike, S. Patrick, S. Cobey, and J. Gerardin. 2021. Geographic and demo-graphic heterogeneity of SARS-CoV-2 diagnostic testing in Illinois, USA, March to December 2020. Working paper (2021).

[16] S. Hsiang et al. 2020. The effect of large-scale anti-contagion policies on the coronavirus (COVID-19) pandemic. Nature (2020).

[17] J. S. Jia et al. 2020. Population flow drives spatio-temporal distribution of COVID-19 in China. Nature (2020).

[18] O. Karin et al. 2020. Adaptive cyclic exit strategies from lockdown to suppress COVID-19 and allow economic activity. medRxiv (2020).

[19] A. King et al. 2015. Avoidable errors in the modelling of outbreaks of emerging pathogens, with special reference to Ebola. Proceedings of the Royal Society B: Biological Sciences 282, 1806 (2015), 20150347.

[20] S. Lai et al. 2020. Effect of non-pharmaceutical interventions to contain COVID-19 in China. Nature (2020).

[21] R. Li et al. 2020. Substantial undocumented infection facilitates the rapid dissem-ination of novel coronavirus (SARS-CoV2). Science 368, 6490 (2020), 489–493.

[22] M. Marathe and N. Ramakrishnan. 2013. Recent Advances in Computational Epidemiology. IEEE Intelligent Systems 28, 4 (2013), 96–101.

[23] M. Marathe and A. Vullikanti. 2013. Computational Epidemiology. Commun. ACM 56, 7 (July 2013), 88–96. https://doi.org/10.1145/2483852.2483871

[24] G. Meyerowitz-Katz and L. Merone. 2020. A systematic review and meta-analysis of published research data on COVID-19 infection fatality rates. International Journal of Infectious Diseases 101 (2020), 138–148.

[25] L. Meyers, N. Dimitrov, and S. Goll. [n.d.]. DiCon: Disease Control System. Available at http://www.bio.utexas.edu/research/meyers/dicon/.

[26] University of Pittsburgh. [n.d.]. FRED – Framework for Reconstructing Epidemi-ological Dynamics. Available at http://fred.publichealth.pitt.edu/tutorials.

[27] University of Pittsburgh. [n.d.]. The Geospatial Area and Information Analyzer (GAIA). Available at http://midas.pitt.edu/gaia.

[28] A. Peddireddy et al. 2020. From 5Vs to 6Cs: Operationalizing Epidemic Data Management with COVID-19 Surveillance. medRxiv (2020).

[29] S. Pei and J. Shaman. 2020. Initial Simulation of SARS-CoV2 Spread and Intervention Effects in the Continental US. medRxiv (2020).

[30] R. Scott and B. Stewart. 2020. Coronavirus cases in the nation’s capital reveal a tale of two cities. ABC News (2020).

[31] S. Thorve et al. 2018. EpiViewer: an epidemiological application for exploring time series data. BMC bioinformatics 19, 1 (2018), 1–10.

[32] W. Van den Broeck et al. 2011. The GLEaMviz computational tool, a publicly available software to explore realistic epidemic spreading scenarios at the global scale. BMC infectious diseases 11, 1 (2011), 1–14.

[33] R. Verity et al. 2020. Estimates of the severity of coronavirus disease 2019: a model-based analysis. The Lancet 20, 6 (2020), 669–677.

[34] C. Wilson. 2020. These Graphs Show How COVID-19 Is Ravaging New York City’s Low-Income Neighborhoods. Time (2020).

[35] B. Wissel et al. 2020. An Interactive Online Dashboard for Tracking COVID-19 in U.S. Counties, Cities, and States in Real Time. JAMIA 27 (04 2020).

[36] Yelp. 2020. Yelp: Local Economic Impact Report. (2020).

